# Cross-Border Socio-Economic Dynamics, community vulnerabilities and the Threat of Cholera Resurgence in Uganda: Insights from Kasese, Hoima, and Kikuube districts

**DOI:** 10.1101/2025.10.13.25337867

**Authors:** Nathan Tumuhamye, Roy William Mayega, Godfrey Bwire, Freddie Ssengooba, Simon Kasasa, Lynn M. Atuyambe

**Author notes:** Corresponding Author: Nathan Tumuhamye. Competing interests statement: No competing interests exist.

## Abstract

**Background:** Cholera remains a global public health threat, causing an estimated 1.3–4 million cases and up to 143,000 deaths annually, mostly in low- and middle-income countries with poor water, sanitation, and hygiene (WASH) systems. In sub-Saharan Africa, outbreaks are driven by poverty, displacement, climate change, and weak health systems. In Uganda, border districts such as Kasese, Kikuube, and Hoima remain vulnerable despite no reported cases since 2018 following vaccination campaigns. Socio-economic activities like fishing, cross-border trade, and seasonal farming continue to heighten exposure. These districts exemplify how livelihoods, mobility, and inadequate infrastructure converge to sustain cholera risks and threaten resurgence even after periods of apparent control.

**Methods:** A qualitative study was conducted in the three districts of Hoima, Kikuube and Kasese in the Albertine region to explore the socio-economic and structural drivers of vulnerability to cholera resurgence. Data were collected through Focus Group Discussions (FGDs), Key Informant Interviews (KIIs), In-Depth Interviews (IDIs), and direct observation. Participants aged 18–77 years included community members, caregivers, health workers, and district officials. Nine FGDs, eleven KIIs, and ten IDIs were conducted. Analysis followed manifest and latent content approaches using open, axial, and selective coding, complemented by ChatGPT 4.5 for verification of meaning units.

**Results:** Inadequate sanitation (open defecation and lack of toilets), unsafe drinking water, seasonal farming settlements without latrines, cross-border movements from the Democratic Republic of Congo (DRC), crowded or makeshift housing, and weak surveillance were identified as underlying vulnerabilities that erode the benefits of vaccination hence drugging these districts to a possible cholera resurgence.

**Conclusion:** The drivers of cholera vulnerability are deeply interconnected: Cholera vulnerability in these districts is rooted in systemic inequities in sanitation, water access, and health infrastructure. Strengthening WASH systems, promoting hygiene behavior change, and improving cross-border surveillance are essential to achieving sustainable cholera elimination and resilience.

**Synopsis:** Cholera, a preventable yet deadly diarrheal disease, continues to threaten millions globally, especially in regions where access to clean water and sanitation is limited. This study, conducted in Uganda’s border districts of Kasese, Hoima, and Kikuube, explored how daily livelihoods such as fishing, farming, and cross-border trade intersect with environmental and social conditions to increase vulnerability to cholera resurgence. Through community discussions and interviews with health workers, local leaders, and residents, the research revealed how open defecation, unsafe water, migration, and poor housing sustain the risk of future outbreaks even after vaccination efforts. The findings provide crucial insights into how human behavior, ecology, and health systems interact, highlighting that preventing cholera is not only a medical issue but a social and environmental challenge. By understanding these links, the study contributes to global health and life sciences knowledge on infectious disease resilience, emphasizing the need for integrated interventions that combine water safety, community education, and stronger health infrastructure.

## Introduction

According to the World Health Organization (WHO), Africa remains one of the most affected continents, with recurrent outbreaks reported in countries such as Nigeria, the Democratic Republic of Congo (DRC), and Mozambique [1]. The cyclical nature of cholera outbreaks in these regions is often linked to the rainy season, which exacerbates the contamination of water supplies, thus heightening the transmission risk [2].

Cholera is an acute diarrheal disease caused by Vibrio cholera, primarily transmitted via contaminated food or faeces. Outbreaks have frequently occurred in Uganda, especially in regions with poor sanitation and high population movement [3]. Over the past 50 years, the border districts of Kasese, Hoima, Kikuube, and Kibale have been identified as cholera hotspots, with a notable increase in cases during the 1990s until 2017 [4, 5]. These areas have been referred to as cholera epicentres [6–8]. Significant outbreaks were recorded from 1997 to 2017, along with sporadic cases reported until 2022 [8, 9]. Since 2017, Kasese has not seen new cholera cases, indicating effective control measures, and there have been no cases reported in Greater Hoima since 2022 [8, 10–12].

Kasese and greater Hoima districts in present with distinct economic conditions and environmental challenges that influence public health dynamics, including the risk of cholera resurgence. Both districts are located in the Albertine region of Uganda. The Albertine region suffers recurrent floods and landslides due to heavy rains. The districts are heavily involved in Agricultural farming and fishing along Lakes Edward, George and Albert. In the greater Hoima (Hoima and Kikuube districts), Agriculture remains the dominant economic activity, with crops such as cassava, maize, and sweet potatoes being widely cultivated [13]. Fishing, particularly on Lake Albert, also plays a significant role in the local economy [14]. Additionally, Hoima’s proximity to oil exploration activities introduces both economic opportunities and environmental concerns, potentially influencing public health outcomes [15]. Kasese District on the other hand is situated in southwestern Uganda, bordered by the Democratic Republic of Congo and characterized by a diverse landscape encompassing the Rwenzori Mountains and Lake Edward. The district’s economy is primarily agrarian, with a significant portion of the population engaged in subsistence farming. Key crops include maize, beans, and cassava, while livestock farming is also prevalent. The presence of national parks, such as Queen Elizabeth National Park and Rwenzori Mountains National Park, contributes to the local economy through tourism. However, the district faces environmental challenges, including deforestation, which has led to increased risks of floods, landslides, and heatwaves. These environmental stressors, coupled with rapid population growth and unregulated timber harvesting, exacerbate the vulnerability of communities to public health threats like cholera [16].

However, the cessation of cases does not mean the risk is gone. Border districts like Kasese, Hoima and Kikuube remain vulnerable due to ongoing socio-economic and environmental conditions. Cholera bacteria can be reintroduced from neighbouring DRC, a country with continuing cholera issues or emerge when local hygiene practices lapse. Between March and April 2025, a qualitative study was conducted in the three border districts. We explored the interplay between socio-economic behaviors, mobility, environmental vulnerabilities, and systemic health challenges that together sustain the risk of cholera resurgence in Uganda’s high-risk border districts. We focused on economic activities, sanitation practices, land use and settlement patterns, migration and cross-border interactions, community health habits, gaps in healthcare and surveillance and related risks that can lead to a resurgence.

## Methods

### Study design and study area

A qualitative study was conducted in Hoima, Kasese and Kikuube districts. The three districts are along the Albertine region bordering the Democratic Republic of Congo. The Albertine region lies along the Lake Albert and River Nile in the mid-western end of the country, and the region comprises the following districts: Nebbi and Zombo in the north, greater Hoima (Hoima and Kikuube) districts in the west and Kasese district in the southwest. The region suffers recurrent floods and landslides due to heavy rains [17]. These have far-reaching impacts on the affected areas, escalating recurrent epidemics of cholera, malaria, and viral haemorrhagic fevers. In Kasese District, the study was conducted in Mpondwe–Lhubiriha Town Council and Nyakiyumbu Sub-County; in Hoima District, Kigorobya and Buresuka Sub-counties were selected; and in Kikuube District, Kabyooya and Kyangwali Sub-counties were included. A mix of qualitative methods including Focus Group Discussions (FGDs), Key Informant Interviews (KIIs), and In-Depth Interviews (IDIs) was employed to capture perspectives across different levels of the health system. FGDs explored community experiences, behaviors, and perceptions of the health system’s drivers of while KIIs offered expert insights from district health officials and administrators, highlighting systemic, underlying gaps in cholera response. On the other hand, IDIs provided detailed accounts from cholera survivors and caregivers, uncovering personal realities and sensitive issues that may not surface in group discussions.

### Study Participants

The study engaged a diverse group of participants to capture perspectives from different layers of the community and health system. Focus Group Discussions (FGDs) included eight to ten participants representing at-risk or affected populations: adult men and women, youth (both male and female), and elderly persons who provided historical context and traditional practices. Caregivers who had directly cared for cholera patients were included, alongside local health workers such as Village Health Team (VHT) members trained in WASH and nurses or health facility staff from affected areas. Community leaders and influencers specifically representatives of the local council leaders, religious leaders, and teachers were also represented, as well as vulnerable groups such as refugees where relevant. On the other hand, Key Informant Interviews (KIIs) were conducted with district-level and institutional actors. These included District Health Officers, District Health Inspectors or Water Officers, health facility in-charges (particularly from Health Centre IIIs and IVs) who oversee case management and referral systems, and surveillance focal persons. Representatives from NGOs and CBOs engaged in water, sanitation, and hygiene (WASH) activities were also included. The In depth interview participants were either survivors of cholera or caregivers of cholera patients

### Data collection procedures

Data was collected between 14-25 April 2025 in all the three study districts. In the same period, all study participants were recruited and interviewed. Prior to data collection, ten (10) research assistants were selected with a minimum qualification of a bachelors degree and with experience in qualitative data collection. The experience as limited to at least have participated in for qualitative studies. Research assistants were recruited from the established pool at the Makerere University School of Public Health (MakSPH), which has been developed over time through various research projects and includes individuals with diverse language competencies. From this pool, 15 assistants for Greater Hoima and 10 for Kasese were initially shortlisted and invited for interviews. Following the selection process, six were chosen for Greater Hoima and four for Kasese. These selected assistants underwent a four-day training led by Author 1 (NT), which included a one-day pre-test of the study tools in Makerere 3 Parish an area with a history of recurrent cholera outbreaks. After training, the teams were deployed to the field for data collection.

Pre-study visits led by NT and one of the research assistants in each district were conducted in the study districts. This pre-visit involved meetings with the District Health Officers (DHOs), the Chief Administrative Officers (CAOs) and the Resident District Commissioners (RDCs) in each district. The districts involved a presentation about the study, the study methods, the selected sub counties and the study participants categories. For the meetings with the District Health Officers, we also identified the respondents for the Key informant interviews and the community mobilizers who were requested to identify the categories of the study participants in the FGDs and IDIs. After these engagements, the district officials endorsed the introductory letters by signing and acknowledging these meetings. Copies of the acknowledgement letters were then provided to the Research Assistants to move with while in the field. The introductory letters were presented to the village chairpersons before any interviews were conducted.

The District Health Officers supported the process by providing contacts for key informants and, in some cases, assisting in scheduling interviews. Using the contacts of Health Inspectors shared by the District Health Officers, the study team coordinated with them to identify suitable FGD participants according to the predefined categories, select appropriate venues, and arrange interviews with in-depth interview participants. At each sub-county, the Health Inspectors shared schedules for FGDs and IDIs with the study team, who then implemented them as planned. In each sub-county, 2–3 FGDs were conducted. All In-Depth Interviews (IDIs) were conducted at the household level, with data collection teams comprising an interviewer and a note-taker visiting each household. Key Informant Interviews (KIIs) were carried out in district offices for officials and in health facilities for facility in-charges. Focus Group Discussions (FGDs) were held either at health facilities or within community settings, as guided by Health Inspectors. Each FGD participant received a transport refund of Uganda Shillings 20,000 (approximately USD 5), as approved by the Ethical Review Board.

Each session (FGD, KII, or IDI) was facilitated by two research assistants (an interviewer/facilitator and a note-taker). Verbal consent was obtained from all participants after the study objectives, risks, and benefits were explained. Discussions were guided by translated interview tools in the local dialects, Runyoro or Rukiga (for Banyoro and Bakiga), Kiswahili (for Lugbara communities) in Hoima and Kikuube, and Lhukonzo in Kasese. To ensure confidentiality, FGD participants were assigned numbers (1–10), which they used instead of names during discussions, with all contributions recorded under these identifiers. Note-takers captured detailed field notes, supplemented by audio recordings. On average, FGDs lasted about 90 minutes, while IDIs and KIIs each lasted approximately 60 minutes.

### Data collection tools and variables

In this study, the FGD, KII and IDI guides with generic questions were used. The guides explored three generic themes

1. Main shock (cholera outbreak), its frequency, primary and secondary effects in the target communities (Risks)
2. Factors that make health system vulnerable to the cholera outbreak (Vulnerability factors)
3. Factors that make the health system fail to permanently resist, reduce or eradicate their vulnerability factors (drivers of vulnerability)

### Data analysis

The audio tape recordings were transcribed verbatim by selected research assistants fluent in the local languages. The first (NT) and last authors (RWM) developed a coding scheme following multiple readings of the transcripts. The coding scheme was then shared with the three research assistants who supported the data analysis. Analysis was done in two stages, first, the manifest content analysis followed by latent content analysis. Transcripts generated from the data were read and re-read by three research officers, who then assigned codes and came up with a coding structure (Open coding). Data meaning units were then aligned under their respective codes in excel. This was followed by axial and selective coding to develop higher codes and sub-themes. Sub-themes were reviewed by the three officers and NT further to develop overarching themes. Chat GPT 4.5 was also used for supplementary analysis especially in verifying the meaning units generated by the three research officers.

### Ethical considerations

Ethical approval was obtained from the Makerere University School of Public Health Research and Ethics Committee (SPH-2024-675). Prior to any of the interviews, including group discussions, a consent form was read out to the participants outlining the goal of the study, the benefits, risks and confidentiality clauses. The study participants were then given a chance to ask questions. After the explanations by the Research assistants, the participants in the IDI and KII were asked to sign two copies of the consent form each of which one copy was retained by each study participant. The forms were also signed by the research assistant on behalf of the study team. For FGDs, group consent was sought using the same processes above. However, this was confirmed by signing the FGD participants attendance lists which include some sociodemographic information like age, gender, education levels and religion.

## Results

A total of eighty-seven (87) participants were interviewed, with an average of 9 people per FGD. A total of 10 FGDs, 13 KIIs and nine (9) IDIs were conducted in the three districts. Cholera thrives in environments where water and sanitation are compromised and can spread rapidly among mobile or dense populations. Inadequate sanitation (open defecation and lack of toilets), unsafe drinking water, seasonal farming settlements without latrines, cross-border movements from the Democratic Republic of Congo (DRC), crowded or makeshift housing, and weak surveillance were identified as underlying vulnerabilities that erode the benefits of vaccination hence drugging these districts to a resurgence. These underlying factors were linked to socio economic activities that define the daily livelihoods of the communities.

### Socioeconomic dynamics exacerbated by Poor Sanitation Practices (Open Defecation) especially within the large farms and in the fishing communities

Socio-economic dynamics strongly shape community vulnerability to cholera, and this was expressed with nuance across FGDs, KIIs, and IDIs. In focus group discussions, poverty and livelihood pressures were repeatedly identified as underlying risks. Poor sanitation practices, particularly open defecation, was reported to be a key driver of cholera outbreaks especially in farming settlements and fishing communities. FGD participants fishing villages like Kibiro in Hoima and Kaiso in Kikuube and farming settlements in Katholhu Kasese district consistently associated cholera outbreaks with poor sanitation and widespread open defecation. In Kibiro, FGD participants explained that fishermen often defecate directly into the lake, contaminating the same water used for drinking and cooking. They also described how seasonal floods break down pit latrines, spreading faecal matter across compounds, gardens, and food markets. In Kasese particularly Katholhu, farming practices were highlighted as key underlying driver of cholera outbreak. It was reported that in farm settings, migrant laborers without access to toilets practice open defecation in the fields, increasing risks of transmission.

> *“flooding brings dirty water from all over, and because the place is low-lying, floods break latrines… that’s how people got sick” FGD participant, Katholhu, Kasese district*

> *“it’s true cholera came because people didn’t have latrines, people were coming from farms, especially in Katholu side, where most people farm from, but they come with their food to the farm, and yet they don’t have any latrines to use. Equally, whenever they bring their food and they want to use the latrines, they find that they don’t have anywhere to do this business. In the farms, people have no water and toilets, and normally people eat yet their hands are contaminated, and that’s why they normally get infected. we have requested owners of land during sensitization to put toilets on these farms, but people don’t heed to this” FGD participant, Katholhu, Kasese district*

> *“these migrants that come through the lake always settle on the lake shores with no latrines and will be practicing open defecation exposing our water source the lake to contamination” FGD participant, Kibiro, Hoima district*

> *“We dispose off the faecal materials into the lake and use the same water for domestic use”. FGD participant, Kibiro, Hoima district.*

These practices were confirmed by key informants. For example In Kasese, it was reported that in large farms, landowners rent out plots to seasonal workers who put up temporary shelters without sanitation facilities, forcing them to defecate in the open. As a practice reported by the district health officials in Kasese, highland communities migrate to lowland farms during the planting season and erect temporary shelters without any toilet facilities (refer to figure 2&3). They end up defecating in gardens, bushes, or open fields. As a result, human waste enters the environment untreated. It was reported that during rainy seasons, faecal matter is washed into streams or ponds that villagers use for water, creating a direct route for cholera transmission

In Hoima and Kikuube especially in the vishing communities, it was reported by district officials that sandy soils and flooding make it difficult to construct lasting pit latrines, while refugee influxes in landing sites overwhelm existing facilities. Officials noted that these conditions contaminate water sources and predispose the communities to outbreaks given the high concentration of households at the lake showers (refer to figure1).

> *“they just put up temporary structures where they settle, but defecate in the open spaces. We are still seeing that happening” KII Kasese district*

> *“I noticed that people were doing open defecation along the water sources in the refugee camp… the feces could go directly into the water” KII Kikuube district*

On the otherhand, a cholera survivor in Nyansenge village, Hoima district, caregivers in Toonya village, Kikuube district and Katholhu village Kasese district offered vivid insights into how open defecation posses a risk to cholera outbreaks. Caregivers described children falling ill after drinking river water contaminated by neighbors whom they perceived to defecate upstream, while others recalled the pain of losing family members due to the lack of latrines that might have exposed them to risks. These IDIs expressed shame, noting that cholera made them feel “like the dirtiest person in the world,” even with those whose homes had toilets because infection often came from surrounding households or farms without facilities.

> *“Those who do open defecation, the rain brings all those feces into the lake which can result in disease outbreaks because of the dirt” Caretaker, Kaiso Tonya, Kikuube.*

> *“Sometimes we would even see faeces flow in the river as you are fetching water… that was what mainly brought us the disease” Caregiver, Nyakiyumbu Kasese district*

> *“Cholera infection shames a lot. I felt as if I was the dirtiest person in the entire world” survivor, Nyakiyumbu, Kasese district*

**Figure 1:**
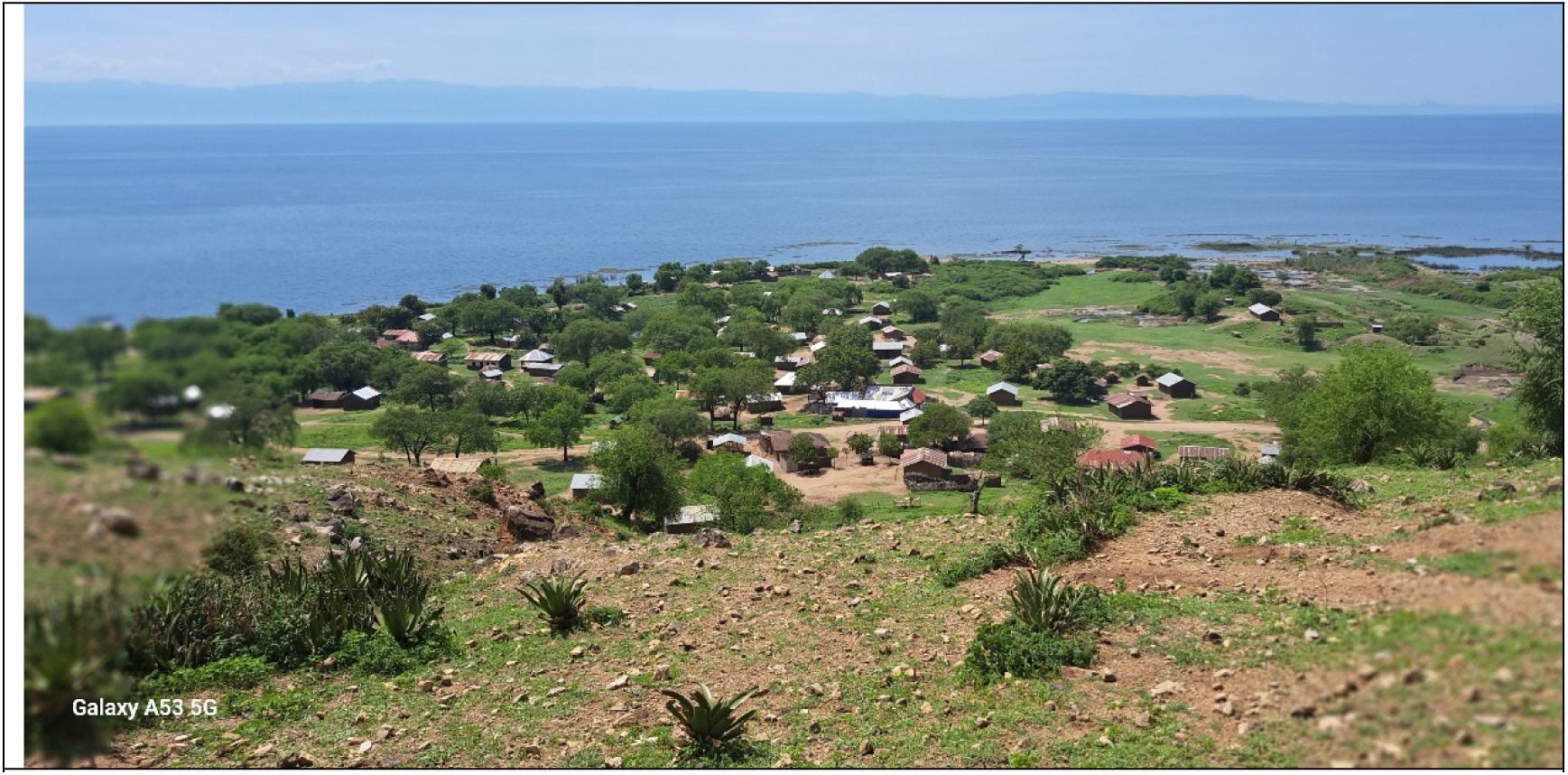
Concentration of the households at Lake Albert, Hoima ditrict

**Figure 2&3:**
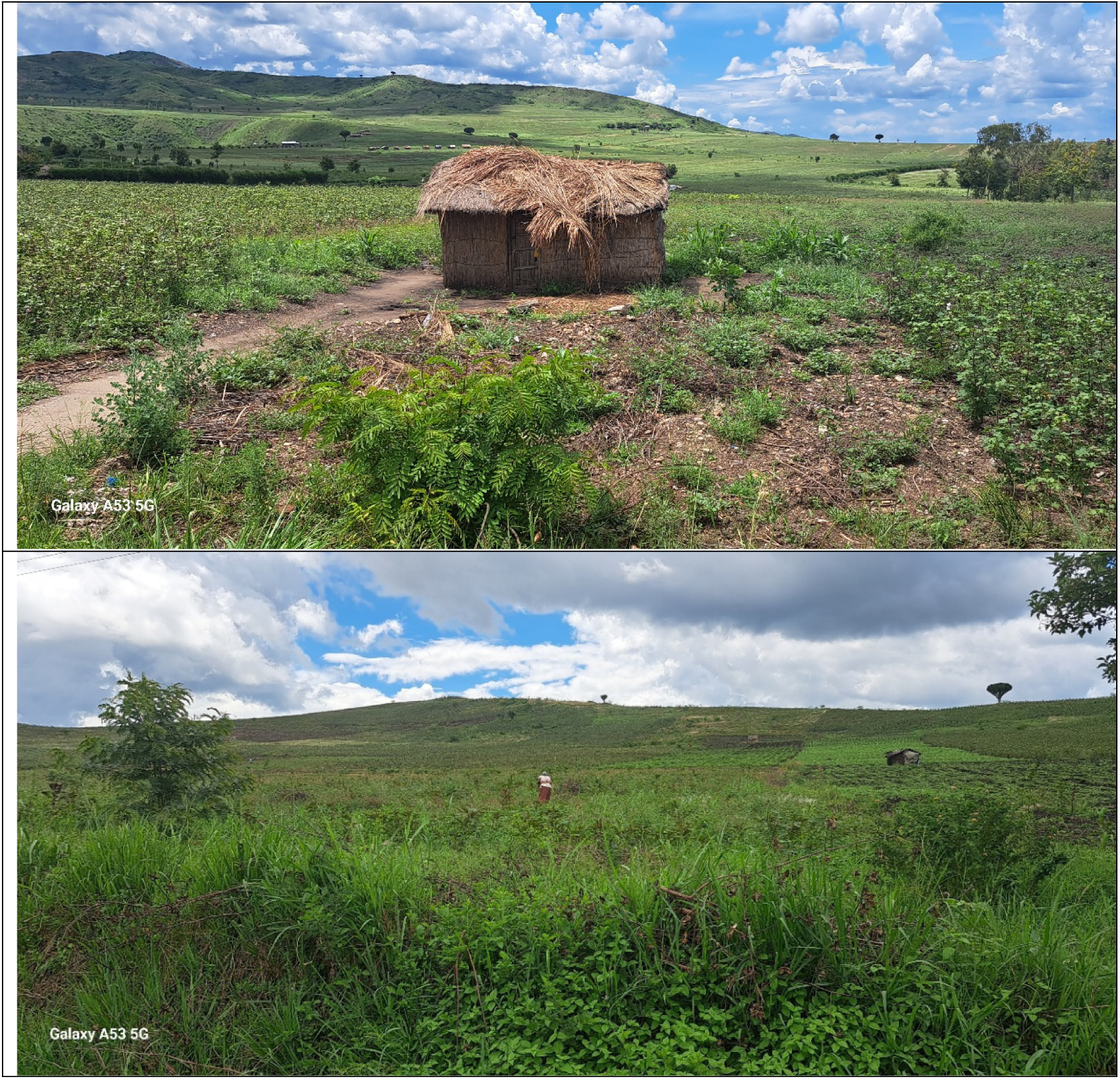
Temporary shelters without latrines in big farms

### Unsafe Water Sources and Practices

It was reported both by study participants that many people both at household level and during farming use untreated, contaminated water for drinking or cooking which poses a major cholera risk. From Focus Group Discussions, community members in Kasese and Hoima stressed how lack of safe water remains an underlying risk to possible cholera outbreaks. In Katholhu Kasese district, FGD participants stressed that “cholera is brought by lack of clean water in the area” and described how floods sweep faeces into drinking water during the rainy season exposing them to a risk of a possible outbreak. Residents in Kibiro echoed this, lamenting that “what we are drinking is contaminated dirty water from the lake where people, children, fishermen defecate”. Individual testimonies reinforced these fears: a caregiver in Tonya, Kikuube district recounted how fishermen *“defecate in the lake and again drink that water… when the water from the lake washes the feces on the shores, we go get that contaminated water and drink it without boiling” IDI caretake, Tonya, Kikuube district*. Similarly, a caretaker in Kasese admitted that before taps were installed, households relied on River Kyanzi, which quickly became unsafe emphasizing that if water from the said river is kept for three days without using it, it will start having a bad smell. It was highlighted that communities in Kasese have historically relied on rivers and streams especially Bukangara River for water for domestic use. In Hoima and Kikuube, it was reported that communities near the lake primarily depend on Lake Albert for drinking water, which they mentioned is significantly polluted due to open defecation by fisher folks and communities around the lake shores.. For example communities in Kibiro during the FGD reported that the only clean water sources are over 6KM away from the residences, and water can only be fetched using motorcycles at a cost of about 0.7USD per 20L jerriycan making people resort to use of contaminated lake water which is affordable

> *“People would fetch water from river Kyanzi… contaminated by people washing upstream. Sometimes we would even see faeces flow in the river as you are fetching water” IDI Caretaker, Nyakiyumbu, Kasese*

> *“One jerrycan is for 2,000 Ugandan shillings but sometimes during the dry season, the jerrycan can go for 2500 shillings… We drink that water even when we know it’s not clean but we have no option.” FGD participant, Kibiro, Hoima*

Key informants framed unsafe water as both an environmental and structural vulnerability. In Kikuube, a key informant from the district health office explained that outbreaks affected everyone equally *“because they are taking the same water, from the same source, unprotected. So, meaning they are eating the same feces”KII, Kikuube district*. The respondent highlighted how shared reliance on contaminated water sources continue to expose the entire populations. In Kasese, another key informant emphasized progress due to government and NGO interventions: *“when they started protecting water sources and people were learning to use safe water, it could be another reason why we have no virus” KII Kasese district*. The respondent emphasized how cholera risk is reduced when safe water systems are established and when communities adopt new behaviors, yet the fragility of these gains in the face of floods, poverty, and weak infrastructure continue to pause a risk for a possible outbreak.

### Migration and Population Mobility

From focus group discussions, communities repeatedly linked migration and mobility to cholera outbreaks. In Kinyansenge, Kikuube district, participants recalled events of 1990s where arriving as migrants from Kigezi in the late 1990s, settled at the landing sites without latrines, and witnessed what they termed *“open defecation everywhere you tried to step”*, which fueled cholera spread. They maintained that even now, migrants on the lake shores do not have latrines and have continued to dispose their faecal products into the lake and on the lake shores. Participants from Kyabikere, Kasese district noted that fishing camps attracted outsiders from other villages and from DRC who defecated in lakes and shared contaminated water, while seasonal farmworkers who moved from either the neighbouring villages in Congo or within other villages in the district often lack latrines. These mobile populations, disconnected from permanent sanitation infrastructure, are seen as major underlying risks of previous and future outbreaks.

From key informant interviews, health workers reinforced this connection, observing that migration patterns made previous outbreak containment difficult. A health inspector in Kasese noted cross-border movement from DRC as a recurring source of cholera importation, stating: *“every outbreak comes from their place then it spreads to us in Uganda here”*. These accounts highlight how both internal and cross-border mobility exacerbate vulnerability.

From in-depth interviews, a survivor from Kyangwali, Kikuube district described being displaced multiple times moving from Kabale to Ankole to Mpocha before settling in Kyangwali and noted how these migrations intersected with cholera exposure especially when the participant settled at the landing site, saying that at landing sites, *“you go to buy fish but it’s like you have bought poison to take home”*. Another caregiver in Kibiro who lost a child recounted the lack of transport or ambulance services in such mobile, hard-to-reach areas, leaving her to rely on ORS at home while her baby died within hours. Together, these perspectives show that migration and mobility create a cycle of vulnerability: mobile groups often lack stable sanitation, introduce infections across communities and borders, and face delayed access to treatment, making cholera outbreaks more severe and widespread.

> *“Much of the origin of the cases we get… they are always from Congo. People cross over and easily transmit the disease” KII, Kasese*

> *“The other reason is that they don’t have permanent settlement… they keep moving, now telling him to construct a toilet for the 3 months they are in the area, they find it hard.” FGD participant, Kibiro, Hoima*

### Housing and Living Conditions

Related to land use and migration is the issue of housing quality in cholera-prone areas From FGDs, communities highlighted how poor housing and settlement patterns made them more vulnerable to cholera. In Katholhu Kasese district, participants described poverty-driven conditions where families lived in houses without latrines, swept waste into compounds, and shared crowded school latrines, creating ripe environments for outbreaks: “cholera is brought about by being dirty… defecating anywhere and then the floods take these pieces all over the compound” FGD participant in Katholhu narrates. In Kyabikere Kasese district, participants lamented that many could not afford proper homes or sanitation facilities, with some still living in temporary shelters or using unsafe latrines that attracted flies into households.

> *“Most people in this community as you see it live in crowded homes with poor or no ventilation and no proper toilets to support many people including mobile communities. It is impossible to keep hygiene standards in our households, and that’s why cholera kept returning before vaccnation.”FGD participants, Kibiro Hoima district*

From KIIs, health officials reinforced these concerns, pointing to structural and land-use problems. The district officials in Kasese noted that seasonal farmers built only makeshift houses and “temporary structures where they settle, but defecate in the open spaces”, with landlords failing to enforce sanitation among tenants on large farms. Similarly, in Kikuube a health inspector explained that collapsible soils and flooding undermined housing and latrine construction, while elderly or impoverished households lacked the capacity to maintain safe facilities. Such fragile housing environments perpetuate open defecation and contamination making the communities and households vulnerable. For example, a caregiver in Tonya observed that houses built near the lakeshore were constantly flooded, preventing latrine construction: “*people close to the lake have no way of constructing latrines, during this rainy season before you put a slab it is destroyed*” *IDI Caregiver Tonya, Kikuube district*. She also described how families refused resettlement even when houses were submerged, choosing instead to rebuild precariously close to flood zones. These accounts show how attachment to livelihoods and geography have forced families to remain in hazardous housing conditions, deepening cholera vulnerability.

### Community Health Behaviors and Perceptions

It was reported that because of non-outbreak period following the vaccination, complacency has set in both in Kasese, Hoima and Kikuube. From FGDs, community members expressed that poor hygiene habits and negligence often fueled cholera. In Kibiro, participants admitted that “*some people in our community defecate anywhere and the rain washes it into the lake and that is the same water we drink” FGD participant Kibiro Hoima district*. Others highlighted improvements after sensitization campaigns, such as practicing handwashing, boiling water, and covering food, but also noted persistent gaps where “stubborn” individuals refused to comply even when leaders emphasized hygiene. In Katholhu Kasese district, it was reported that some households stopped treating drinking water as well as allowing children to play in drainage channels following cholera vaccination (OVC), making the population susceptible to a possible outbreak. It was also reported that due to the economic pursuit (parents working), there is a reduction in oversight of hygiene at household exacerbated by cultural beliefs where people believe that cholera is caused by spiritual forces. Similarly, FGD participants in Katholhu credited government vaccination campaigns and behavior change messages for reducing outbreaks, *saying “we now clean our jerricans, wash our utensils, and know that latrines are the main source (faeces) that brings cholera” FGD participant, Kyabikere, Kasese district*.

> *“They have their own beliefs; that a pregnant woman does not go to the toilet… that’s why they don’t construct toilets.” FGD participant, Kibiro Hoima district”.*

From KIIs, health officials described community behaviors as both barriers and opportunities for prevention. A health inspector in Kasese observed that farmers often ate without washing hands and children left at home “just eat anything they come across,” making them highly exposed to cholera. He also noted the critical role of VHTs in reinforcing behavior change, distributing aqua tablets, and mobilizing households for sanitation campaigns. These insights underline how daily routines, food handling, and community negative perceptions towards the preventive methods and cholera modes of transmission undermine the control despite formal interventions. A caregiver in Tonya admitted that *“failure to adopt hand washing practices all the time… has been so difficult for people” and that many still resisted boiling water, treating it “not as a serious matter” IDI Caregiver Tonya Hoima district*. In Katholhu Kasese district, another caregiver stressed that negligence worsened risks: *“one can finish a week without sweeping their compound… peel food without washing her hands… these are the people we live with”*. Survivors like one from Kyangwali also linked cholera directly to lifestyle and hygiene practices at landing sites, where flies from open defecation easily contaminated food.

### Structural Weaknesses in Healthcare and Surveillance

From FGDs, community members emphasized the fragility of local health systems in responding to cholera. In Kyabikere Kasese district, residents admitted their Health Centre II could not manage severe cases: *“for anyone with cholera, we can’t put such a patient here unless we are saying let him die” FGD participant Kyabikere Kasese district*. Others in Kibiro, Hoima district criticized weak enforcement and absent health staff, saying, *“the health workers just sit in offices, they don’t come to us… by the time you hear about cholera, people are already dying” FGD participant Kibiro Hoima district*. These views reflected widespread frustration that facilities and outreach failed making the communities vulnerable.

From KIIs, officials openly acknowledged systemic shortcomings. A Kasese district official confessed, *“we still have a weakness in surveillance, particularly at cross-border points where response was delayed until World Health Organization* intervened” KII Kasese district. In Hoima, a frontline health worker noted that districts often waited for supplies from Kampala: *“we don’t have chlorine or ORS here; everything must come from the Ministry, and by the time it reaches, people have already died” KII_Frontline Health Worker Hoima district*. Another KII in Kikuube pointed out that refugee influxes and porous borders stretched resources beyond capacity further exacerbating vulnerability.

> *“when there is a problem, it is only WHO to give us a situation analysis of Congo – why not get information directly?” KII, Kasese.*

This indicates a gap in cross-border collaboration and communication. Additionally, it was reported that local surveillance is under-resourced mainly with insufficient transport (vehicles or motorbikes) for health workers to investigate cases in remote areas, and limited training or support for village health teams (VHTs) to report cholera signs early. These structural weaknesses mean an outbreak could smoulder undetected or uncontained for longer, increasing its spread.

From IDIs, families gave painful testimony of how these weaknesses translated into loss of lives. A caregiver in Tonya, Hoima district described the impossibility of reaching care during rains: “in this rainy season there is no way you can reach such areas unless you walk” *IDI_Caregiver Tonya Hoima district*. Another mother from Kibiro, who lost her child, recalled, “*I asked them to give me an ambulance, but they said there was none… I remained at home and continued giving her ORS. She passed on within hours” IDI_Caregiver who lost a child.* This caregiver also narrated how to health facilities have submerged in the water near the lakeshores due to flooding and have for the last 10 years been out of use forcing government to construct a third health facility many kilometres away from the lakeshores where people reside. Survivors also described makeshift treatment centers in schools and tents, where overcrowding and lack of drugs left families fearful that

## Discussion

The convergence of the above risk factors related to socio economic activities especially around farming and fishing in border regions like Kasese and Hoima has serious public health implications. We observed continued activities around the lake and large farms despite the gaps in preventive measures. The continued risky activities that are not followed by the preventive practices pose a very high exposure that might result into a possible resurgence of cholera outbreak. If the highlighted issues in the results are not addressed, the districts could see a resurgence of cholera potentially large, fast-spreading outbreaks with high illness and death rates.

Economic activities such as farming and fishing were highlighted as drivers of previous outbreaks in the study districts. It is important to note that the major livelihood activities in the study districts are farming for Kasese, Hoima and Kikuube and fishing for Hoima and Kikube. These daily activities are conducted by mobile communities that establish mobile settlements either in the gardens or at the lake shores. As indicated, most of these mobile shelters do not have sanitary facilities like pit latrines and faecal disposal is done either in the bushes near the farms or in the lake hence exposing the same communities to contaminated water sources originating from the poor faecal disposal practices. These findings are in agreement with other studies that have documented a linkage between livelihood activities and cholera outbreak. For example in Tanzania, it was reported that “Cholera was found to disproportionately impact communities living along the shores of Lake Victoria in Ilemela and Lake Tanganyika in Nkasi, especially fishermen and women involved in fish trading. Key contributing factors to the outbreaks included defecation in shallow and shoreline areas of the lakes, open defecation, bathing or swimming in contaminated water, and improper waste disposal.” [18]. Other studies have reported such activities as key drivers to the outbreak and spread of cholera [19–22]

Farming and fishing livelihoods, while central to the socio-economic survival of communities in Uganda’s Albertine region, have also amplified vulnerability to cholera outbreaks. Fishing communities are often located along unprotected lakeshores where poor sanitation infrastructure and open defecation directly contaminate water sources, a pattern consistent with studies that identify fishing villages as recurrent hotspots of cholera transmission in Uganda and across East Africa [23, 24]. Similarly, large-scale farming attracts seasonal and migrant laborers who erect temporary shelters without latrines, perpetuating open defecation and unsafe water use, conditions highlighted by research as drivers of both endemic and epidemic cholera [2]. Overcrowding in camps, reliance on contaminated rivers for drinking and domestic use, and mobility between rural farms and trading centers create ideal conditions for rapid disease spread. Evidence from previous outbreaks in Kasese and Hoima districts further shows that poverty linked to these economic activities forces households to prioritize livelihoods over health practices, reinforcing cycles of exposure [25]. Thus, while fishing and farming sustain household incomes, their structural and environmental contexts intensify cholera transmission dynamics and hinder long-term control.

Findings indicated an existence of contaminated water sources as reported by respondents in Kasese, Hoima and Kikuube, resulting from the disposal of faecal materials in the same water sources used domestically. This is especially during rainy seasons where flooding exposure the water sources leading to contamination. These findings are in agreement with previous reports that indicated that for example, in past outbreaks, water tests in Kasese’s affected villages showed high faecal coliform counts in both open streams and piped water, indicating faecal contamination [7]. In other studies, it was reported that many households were drinking water without boiling or disinfecting it. For instance, one study during a 2015 cholera outbreak found “94% of cholera cases had drunk water without boiling compared to 79% of healthy controls” [7]. It is also important to note that while improvements have been made in recent years in extending clean water to communities over the last five years where the government protected more water sources and communities started using safe water with a 57% of the population in Kasese for example having clean water sources with a focus on boreholes, gravity flow schemes, and rainwater harvesting systems [26]. Even so, any lapses in water treatment (like shortages of water purification tablets, or assuming piped water is safe when it’s not consistently chlorinated) can allow cholera bacteria to infect families. For example, an investigation of the 2015 outbreak found that the piped water supply was inadequately chlorinated and became re-contaminated along the pipeline and in home storage [12]. Therefore, intensifying the use of Chlorine and UV disinfections should be part of the preventive measures as these have been found to be effective in water purification and safety of drinking water [27, 28].

Cross boarder migrations for individuals seeking economic activities, medical care or even family related migrations were found to be a key driver of cholera outbreak in the study districts. Respondents indicated a strong relationship between cross border migration and previous outbreaks with many index cases of the previous outbreaks reported to be originating from the Democratic Republic of Congo [4, 8, 24, 29, 30]. Studies have documented a clear linkage between migration and the spread of disease outbreaks [31–34]. The 2015 cholera outbreak in Kasese “originated from Bwera Sub-County, a cholera-prone area near the DRC border where another cholera outbreak was ongoing in DRC” [12]. Once the bacteria are introduced, highly mobile groups can disseminate it widely. In border communities, it’s common for families to be split across the two countries e.g. a man might have one household in Uganda and another in DRC, or children go to schools on opposite sides of the border. Daily cross-border commuting for farming or commerce is also routine. This intermingling means infectious diseases like cholera do not respect borders. An illness acquired in one country is often carried into the other. Migrants and refugees fleeing instability in DRC add to this mobility. An outbreak in a cross-border district can easily spread to or from the DRC due to ongoing movement of people. This complicates control efforts, as two countries must coordinate to trace contacts and contain cases. If not quickly contained, cholera could also spread within Uganda. While Uganda has been successful in containing past cholera episodes to limited areas, the persistence of risk factors means there is always a possibility of a more widespread epidemic if multiple hotspots flare up simultaneously. Such a scenario would have grave public health implications nationwide: cholera could hit urban centers or refugee camps in other regions, amplifying the humanitarian impact.

It was also reported that health facilities along the border districts like Bwera Hospital usually treat not only locals but also patients crossing from DRC, often without additional funding or supplies. This was reported to have led to frequent stockouts of essential emergency medicines and IV fluids during outbreaks, hampering effective treatment. These fundings are in line with findings from the comprehensive Facility-Based Assessment for Emergency, Critical and Operative care conducted by the Ministry of Health in 2024, where Bwera Hospital and Hoima Regional Referral Hospital experienced frequent stock-outs, impacting patient care and emergency procedures [35]. Uganda’s funding formulas do not account for these “unbudgeted” populations, creating a chronic resource gap at the border. If a cholera outbreak happens, such shortages could cost lives. Moreover, disease surveillance systems have shortcomings. Surveillance officers in the study districts reported a limited ability to get real-time information from across the border. International protocols often require going through the World Health Organization (WHO) for cross-border data, which delays the flow of information. This challenge is even magnified by inadequate staffing. For example Bwera Hospital expected to handle emergencies like Cholera and other outbreaks has 160 staff against a required 341 which is only 46.9% of the required staffing [35].

## Conclusion and recommendations

Despite progress through vaccination and improved water access in some areas, the threat of cholera resurgence remains high. Mobility across borders, seasonal migrations, environmental hazards like floods, and weak health surveillance create fertile ground for new outbreaks. The resurgence of cholera in Uganda’s border regions is not inevitable, it is a risk that can be managed with proactive, sustained measures. To reduce the persistent threat of cholera and supplement on vaccination programs, interventions in the Albertine region must focus on strengthening WASH infrastructure, cross-border coordination, behavior change, and health system preparedness. Expanding access to safe water and flood-resistant latrines in fishing and farming settlements is critical, alongside provision of water treatment supplies and community enforcement of sanitation bylaws. Cross-border and migration-aware strategies are essential, including joint Uganda–DRC surveillance, sanitation facilities for seasonal workers, and integration of cholera prevention in refugee programs. Sustained hygiene education through VHTs, schools, and cultural institutions should reinforce handwashing, safe food practices, and water treatment, while addressing myths and negligence. Housing and settlement planning must prioritize relocation or upgrading of households in flood-prone areas and require landlords and farmers to provide basic sanitation. At the same time, frontline health facilities should be upgraded with trained staff, cholera kits, and reliable transport for rapid response, supported by stronger local surveillance and cross-border data sharing. Finally, tackling socio-economic drivers is vital by supporting alternative livelihoods and co-designing community-owned solutions to ensure that cholera prevention is both sustainable and equitable.

## Data Availability

This was a qualitative data that was collected through Focus Group Discussions (FGDs), Key Informant Interviews and In-depth interviews. I have attached the the excel data analysis sheet. May require transcripts beyond the analyzed excel sheet, i will be able to share.

NA

## Notes

### Competing Interest Statement

The authors have declared no competing interest.

### Clinical Trial

NA

### Clinical Protocols

NA

### Funding Statement

The author(s) received no specific funding for this work.

### Author Declarations

This study was approved by Makerere University School of Public Health Research and Ethics Committee with approval number (SPH-2024-675).

